# Evaluating risks and benefits of Newer Diabetes medications when Used in Routine care (ENDURe)

**DOI:** 10.1101/2025.02.20.25322631

**Authors:** Michael Colacci, Mack Hurst, Jamie Lee Fritz, Patricia Olar, Prachi Ray, Amol Verma, Fahad Razak, Muhammad Mamdani, Kieran L. Quinn, Laura Rosella, Michael Fralick

**Author notes:** Correspondence: Michael Fralick, MD, PhD, SM, FRCPC, Sinai Health System 60 Murray Street M5T 3L9.

## Abstract

**Introduction:** Sodium glucose co-transporter 2 inhibitors (SGLT2i) and glucagon-like peptide 1 analogues (GLP1a) improve clinical outcomes (e.g., myocardial infarction, stroke, death from CV causes) for adults with type 2 diabetes mellitus. The majority of available data from clinical trials and observational studies are from outpatients. Data on their effectiveness among hospitalized patients, however, are lacking.

**Methods:** We conducted a multicentre, retrospective, cohort study of adults aged 65 years and older with type 2 diabetes mellitus hospitalized between 2017 and 2023 in Ontario. We compared adults newly prescribed an SGLT2i or GLP1a in hospital to adults newly prescribed a DPP4i in hospital. In a sensitivity analysis, new use of sulfonylurea was the comparator. Our primary outcome was the 1-year risk of a composite of all-cause mortality, hospitalization with myocardial infarction, stroke, heart failure, or renal failure. Secondary outcomes included components of the composite, short-term (i.e., 30 day) risk of hypoglycemia, and the 1-year risk of DKA.

**Results:** We identified 6,713 older adults with diabetes who were newly prescribed one of the following medications during an inpatient hospitalization: SGLT2i (N=1520), GLP1 (N=90), DPP4i (N=3726), sulfonylurea (N=1377). Because new use of GLP1 was rare in hospital, we updated our exposure group to SGLT2i alone. Adults who received an SGLT2i were typically younger, and more likely to have heart failure or coronary artery disease compared to adults who received a DPP4i or sulfonylurea. Among adults who received an SGLT2i, at 1-year the primary outcome occurred in 26% compared to 31% who newly received a DPP4i (adjusted hazard ratio [HR] 0.82 95% Confidence Interval [CI] 0.69,0.97). In our sensitivity analysis using sulfonylurea as the comparator group, the hazard ratio for the primary outcome was 0.97 (95% CI 0.80, 1.18). We did not identify an increased risk of DKA or hypoglycemia for SGLT2i compared to DPP4i, though patients receiving an SGLT2i did have a lower rate of 30-day readmission (HR 0.73, 95%CI 0.58-0.92).

**Conclusion:** Among older adults with type 2 diabetes mellitus, newly prescribing an SGLT2i or GLP1 during a hospitalization was uncommon. New use of an SGLT2i was associated with improved outcomes compared to DPP4i but this finding was not robust when new use of a sulfonylurea was the comparator. Future larger studies are needed to provide more definitive results.

## Introduction

Sodium glucose co-transporter 2 inhibitors (SGLT2i) and glucagon like peptide-1 analogues (GLP1a) are among the most effective medications for people living with type 2 diabetes mellitus (T2DM).^1–5^ Both classes of medications reduce a person’s composite risk of myocardial infarction, stroke, and cardiovascular mortality.^1–3,5^ With SGLT2i, the number needed to treat (NNT) to prevent myocardial infarction, stroke, or death from cardiovascular causes is approximately 80 (over 3 years).^1–3^ For adults with T2DM, SGLT2i reduce a person’s risk of end-stage kidney disease (NNT 25, 2 years) and the composite of heart failure or cardiovascular death (NNT 20, 2 years).^6–8^ GLP1a have comparable cardiovascular benefits (NNT 80, ∼2 years), and a recent clinical trial and meta-analysis suggests they may also reduce a person’s risk of renal failure or heart failure.^9,10^

Guidelines now preferentially recommend SGLT2i and GLP1a as second-line agents for adults with T2DM who have established or increased risk of cardiovascular disease.^11,12^ Despite these guidelines, however, the uptake of both classes of medications for older adults into clinical practice has been slow.^13–16^ Slow uptake is common with new medications for chronic diseases. In the case of SGLT2i, this is exacerbated by an important, albeit rare, adverse side effect of the medication: risk of diabetic ketoacidosis (DKA) (number need to harm = 200, 1-year).^17–19^ However, recent studies demonstrate that DKA risk can be estimated based upon insulin use, hemoglobin A1c and prior DKA events ^20^.

Hospitalizations represent a potential important opportunity to initiate guideline-indicated pharmacotherapy. The average age of a patient hospitalized with type 2 diabetes is approximately 70 years, and prior work has shown that half of these patients have cardiovascular disease, heart failure or renal failure.^21^ Data on the effectiveness of SGLT2i or GLP1a started during an inpatient hospitalization are lacking as most studies primarily recruited outpatients. Our primary objective was to estimate the long-term (e.g., heart failure hospitalization, cardiovascular event) outcomes of initiating an SGLT2i or GLP1a during a hospitalization.

## Methods

### Population of Interest and Sampling Methods

We conducted a population-based, active comparator, retrospective cohort study. We obtained our sample from ICES administrative database linked with GEMINI. We included adults over the age of 65 years who were diagnosed with type 2 diabetes and newly prescribed an SGLT2i/GLP1a, or a sulfonylurea/DPP4 inhibitor between April 1^st^ 2017 (following the addition of SGLT2i to Ontario drug formulary) and January 1^st^ 2023 (most recent available data). Patients with type 2 diabetes were identified using the International Classification of Diseases, Ninth Revision (ICD-9) and ICD-10 codes which have a positive predictive value of over 90% based on a recent validation study in GEMINI^22^.

Cohort entry was defined as the date of the first prescription for SGLT2i/GLP1a or sulfonylurea/DPP4 inhibitor during the patient’s hospitalization. We excluded adults with type 1 diabetes mellitus (who aren’t treated with SGLT2i/GLP1a), patients on dialysis (who are not eligible to receive SGLT2i), age > 90 years (because of the strong selection bias against use of SGLT2i or GLP1 in this age group)^23^, seen by palliative care in the last 1-year (as a palliative philosophy of care may affect a person’s likelihood of presenting to hospital and therefore outcome ascertainment) and participants with insufficient baseline healthcare data (i.e., less than 365 days).

ICES is an independent, non-profit research institute whose legal status under Ontario’s health information privacy law allows it to collect and analyze health care and demographic data, without consent, for health system evaluation and improvement. The ICES database provides de-identified longitudinal, individual-level data on patient demographics, healthcare utilization, medical diagnoses, diagnostic tests, clinical procedures, outpatient laboratory results, and pharmacy dispensing of drugs to over 10 million people in Ontario, Canada. The GEMINI database includes administrative data and clinical data for all patients admitted to, or discharged from, the general internal medicine units at the affiliated sites.

## Data collection

Data from ICES included pharmacy medication dispensing (including diabetes and non-diabetes medications), outpatient laboratory results (e.g., hemoglobin A1c, creatinine) and vital status. Clinical details obtained from the hospitalization via GEMINI included demographic information (age, sex), comorbid conditions (using Clinical Classification Software Refined [CCSR] codes, e.g., hypertension, coronary artery disease), inpatient medications (e.g., insulin, metformin), inpatient diagnostic test results (laboratory values), physician orders and hospitalization outcomes (length of stay, and inpatient mortality). The accuracy of GEMINI data (laboratory, radiology, physicians, death, transfers, and transfusion) relative to manual chart review by a trained chart abstractor have previously been evaluated and data fields have an accuracy of above 97%.^11^ These datasets were linked using unique encoded identifiers and analyzed at ICES.

## Exposure and Comparator Groups

The exposure group was defined as adults with type 2 diabetes who were newly prescribed an SGLT2i or a GLP1a in hospital. The main comparator group was adults with type 2 diabetes who were newly prescribed another second-line medication (i.e., DPP4 inhibitor) during their hospital admission. In a sensitivity analysis the comparator group was sulfonylurea to assess the robustness of observed results in our primary analysis. We defined new use as not having received the same class of medication (SGLT2i, GLP1a, DPP4, sulfonylurea) for the past 365 days prior to hospitalization. For example, a patient was defined as a new user of an SGLT2i if they received empagliflozin in hospital but had not received another SGLT2i (empagliflozin, canagliflozin, dapagliflozin, etc.), in the 365 days prior to hospitalization.

A new user of an SGLT2i was then compared to an adult newly started on a DPP4i. We excluded patients who were newly started on both an exposure and a comparator during the same hospitalization.

## Study Outcomes

The primary outcome was a composite of myocardial infarction, stroke, heart failure hospitalization, renal failure or death over a 365 day period. As a secondary analysis, we evaluated the individual components of the composite outcome. Myocardial infarction, stroke, and heart failure hospitalization were defined using the validated ICD-10 codes, which have a PPV exceeding 85%.^24–26^ Renal failure was defined using the ICES dictionary^27^. Death was evaluated using ICES vital status data. Safety outcomes included hospitalization with DKA and severe hypoglycemia. Hospitalization with DKA was defined using ICD-10 codes. These codes were recently validated within GEMINI hospitals by our study team and had a positive predictive value of 70% and negative predictive value above 99%.^28^ Severe hypoglycemia was defined as a glucose less than 2.2 mmol/L and assessed on the index admission and up to 30 days following the date of discharge. We did not have access to outpatient glucose measurements performed within the patient’s home or at visits with their family doctor or endocrinologist.

## Statistical Analysis

A Cox proportional hazards model was used to examine the association between the exposure and outcome while adjusting for confounding. Covariates included in the model were based upon prior literature^29,30^ and our teams collective clinical judgment, and included: age, sex, comorbidities (hyperlipidemia, coronary artery disease, chronic obstructive pulmonary disease, cirrhosis, chronic kidney disease, cerebrovascular disease, heart failure and dementia), medication use (metformin, insulin, GLP1, diuretics, antiplatelets, anticoagulants, statins), and laboratory values (creatinine and hemoglobin) prior to starting on the medication of interest. The lab values included both outpatient and inpatient values. We used an intention to treat approach (i.e., patients who started on an SGLT2i were analyzed accordingly regardless of whether they switched to the comparator during follow-up). We censored patients at death or the end of the 365-day follow up period. Statistical analysis was completed using SAS Enterprise Guide version 8.3.

## RESULTS

Table 1 reports the baseline characteristics of patients for each class of medication. In general, adults who received an SGLT2i were typically younger than other groups, 38.8% were female, 36.5% had heart failure, 50.5% were receiving metformin, and the median hemoglobin A1c was 7.2% (Interquartile Range [IQR] 6.5-8.3%). In comparison to other groups, those who received a DPP4 inhibitor were typically older, 42.8% were female, 19.6% had heart failure, 46.9% were receiving metformin, and median hemoglobin A1c was 7.4% (IQR 6.6-8.8%).

**Table 1.** Baseline characteristics of patients.

|  | Total | Med Class |  |  |  |
| --- | --- | --- | --- | --- | --- |
|  |  | sglt2 | glp1 | dpp4 | sulfonylurea |
|  |  | N=1520 | N=90 | N=3726 | N=1377 |
| **Total** | 100% | 22.6% | <6.7% | 55.5% | 20.5% |
| **Demographics** |  |  |  |  |  |
| **Sex** |  |  |  |  |  |
| Female | 41.6% | 38.8% | 45.6% | 42.8% | 41.2% |
| **Age** |  |  |  |  |  |
| 65-69 | 22.6% | 23.4% | 34.4% | 21.0% | 25.0% |
| 70-74 | 22.6% | 25.7% | 35.6% | 20.9% | 22.9% |
| 75-79 | 20.5% | 22.8% | 18.9% | 19.5% | 20.8% |
| 80-84 | N/A | 16.3% | <6.7% | 20.1% | 17.4% |
| 85-89 | N/A | 11.8% | <6.7% | 18.5% | 13.9% |
| **Comorbid conditions** |  |  |  |  |  |
| Hypertension | 24.1% | 25.4% | 22.2% | 23.9% | 23.2% |
| Coronary Artery Disease | 12.9% | 18.8% | 12.2% | 11.1% | 11.3% |
| Renal Failure | 11.7% | 9.7% | 7.8% | 13.6% | 9.3% |
| Stroke or TIA | 3.5% | 2.8% | - | 4.1% | 2.9% |
| Heart Failure | 23.4% | 36.5% | 16.7% | 19.6% | 19.4% |
| Dementia | N/A | 6.3% | <6.7% | 12.6% | 11.5% |
| **Diabetes Medications** |  |  |  |  |  |
| SGLT2 Inhibitor | 9.5% | - | 27.8% | 10.6% | 16.0% |
| GLP1 analogue | 2.6% | 3.0% | - | 2.7% | 2.3% |
| Dipeptidyl Peptidase 4 Inhibitor | 21.8% | 40.5% | 22.2% | - | 60.1% |
| Sulfonylurea | 19.2% | 29.5% | 22.2% | 22.0% | - |
| Metformin | 46.0% | 50.5% | 53.3% | 46.9% | 38.0% |
| Insulin (long,short, intermediate acting) | 26.6% | 27.0% | 66.7% | 27.2% | 22.0% |
| **Non-Diabetes Medications** |  |  |  |  |  |
| Anti-hypertensives | 79.6% | 84.4% | 86.7% | 78.0% | 78.3% |
| Diuretics | 26.9% | 31.3% | 27.8% | 26.9% | 22.1% |
| Antiplatelet | 14.5% | 16.4% | 24.4% | 13.6% | 14.3% |
| Anticoagulant | 20.1% | 26.0% | 18.9% | 18.8% | 17.0% |
| Statin | 70.7% | 75.2% | 81.1% | 68.1% | 72.3% |
| **Lab Values** |  |  |  |  |  |
| **Creatinine (umol/L)** |  |  |  |  |  |
| % of patients that had lab values | 98.9% | 98.9% | 98.9% | 98.9% | 98.9% |
| 25th Percentile | 79 | 75 | 71 | 82 | 78 |
| Median | 105 | 94 | 102 | 116 | 102 |
| 75th Percentile | 152 | 120 | 150 | 175.5 | 142 |
| **Hemoglobin A1C (proportion)** |  |  |  |  |  |
| % of patients that had lab values | 86.8% | 88.7% | 96.7% | 85.8% | 86.9% |
| 25th Percentile | 6.6 | 6.5 | 7 | 6.6 | 6.8 |
| Median | 7.4 | 7.2 | 7.7 | 7.4 | 7.6 |
| 75th Percentile | 8.7 | 8.3 | 8.7 | 8.8 | 9 |

### Primary outcome

Prior to adjustment, the primary composite outcome (myocardial infarction, ischemic stroke, heart failure hospitalization, renal failure hospitalization, or death) occurred in 859 (31%) of patients who received a DPP4i and 332 (25.5%) who received an SGLT2i. After adjustment, this corresponded to a hazard ratio (HR) of 0.82 (95% Confidence Interval [CI] 0.69-0.97). Prior to adjustment, the primary composite outcome occurred in 274 (26%) of patients who received sulfonylurea and 332 (25.5%) who received an SGLT2 inhibitor (Figure 2). After adjustment, this corresponded to a HR of 0.97 (95%CI 0.80- 1.18) . The components of the composite are reported in Table 2. The most common event contributing to the primary outcome was death, which occurred for 608 (22.0%) of patients who received a DPP4i, 179 (17.0%) who received a sulfonylurea and 203 (15.6%) who received an SGLT2i.

**Figure 1.**
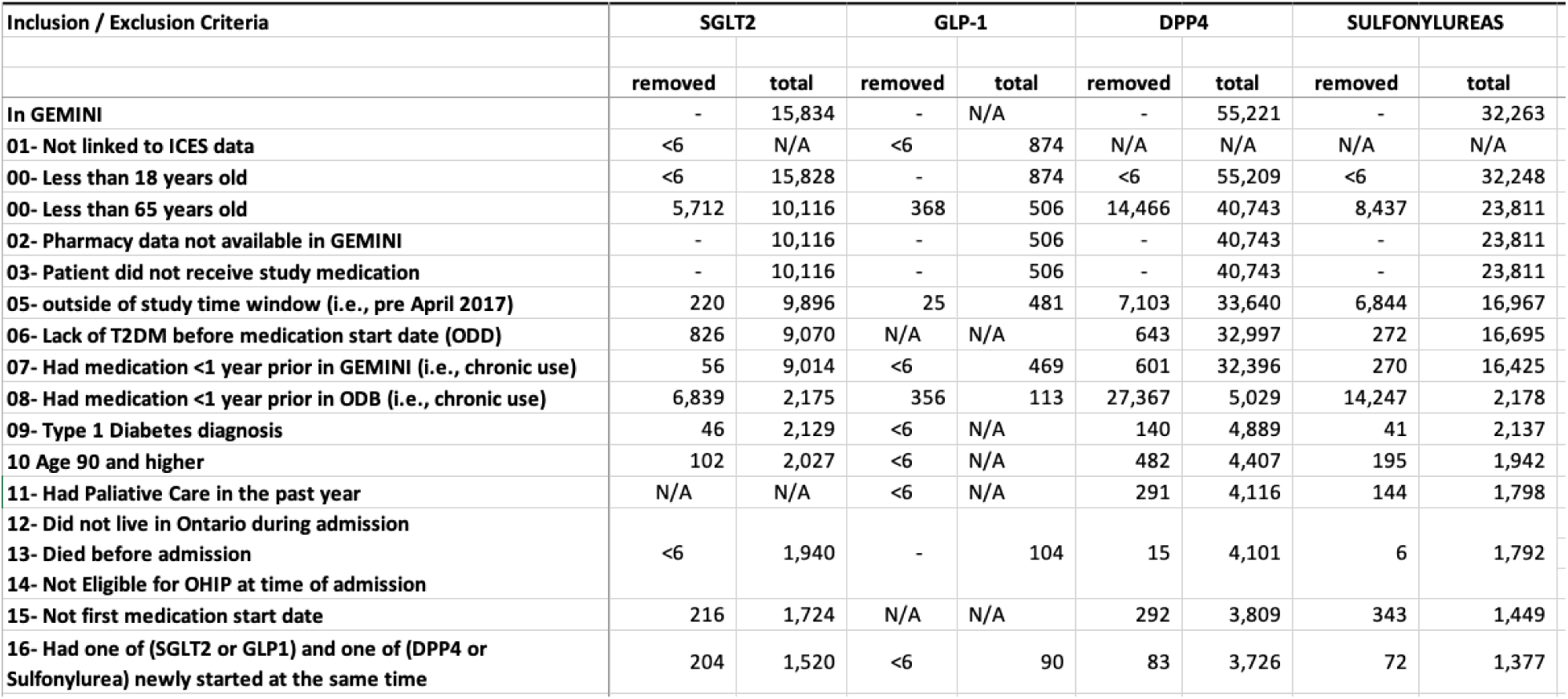
An overview of the proposed study design

**Figure 2.**
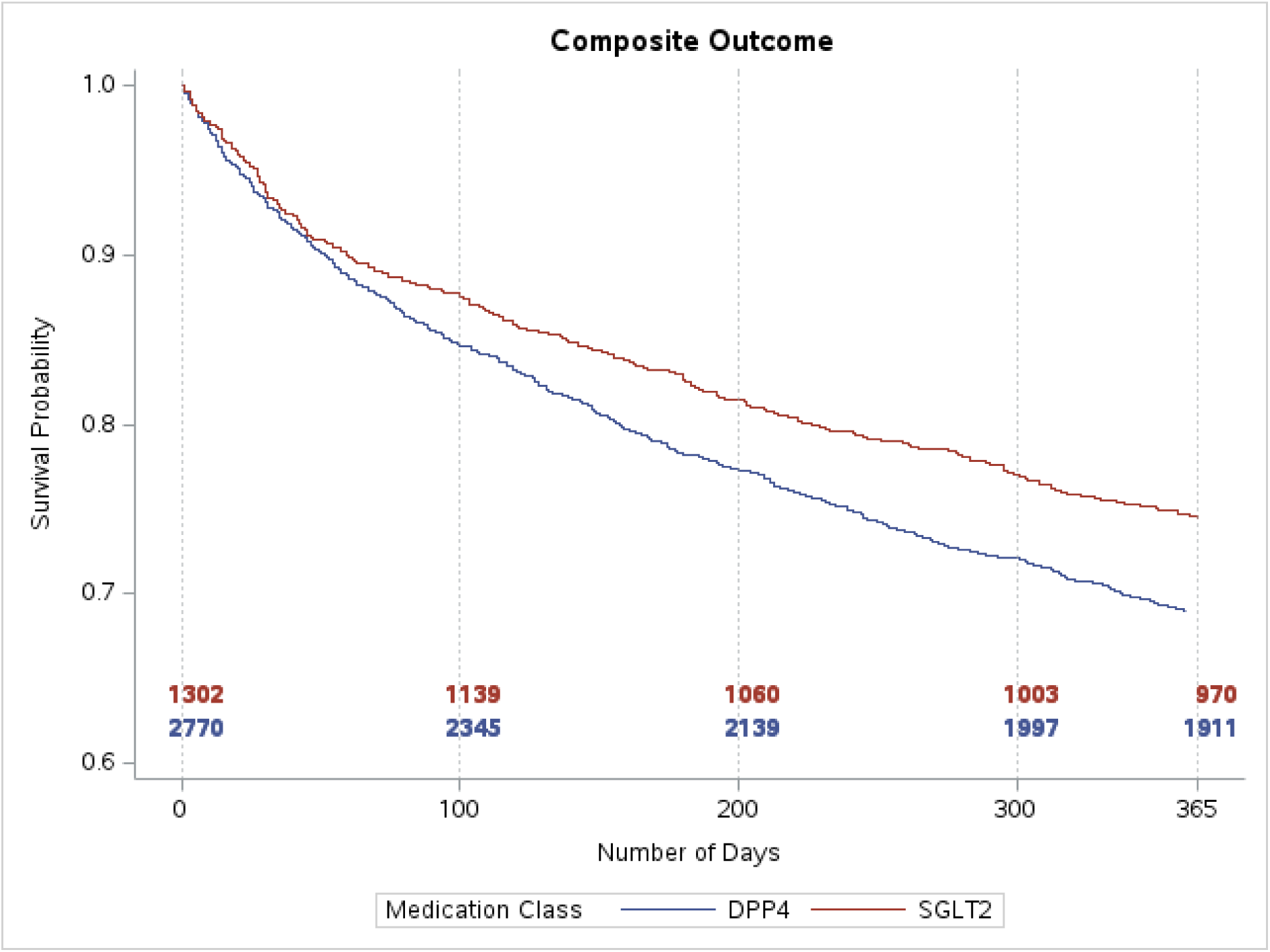
Kaplan meier curve for primary outcome Primary outcome was a composite of cardiovascular events, renal failure, or heart failure hospitalization. DPP4 = Dipeptidyl peptidase 4 inhibitor SGLT2i = sodium glucose cotransporter 2 inhibitor

**Table 2.** Primary and secondary outcomes for SGLT2i compared to DPP4i.

| Outcome | SGLT2i | DPP4 | Adjusted Hazard Ratio<br>(95% Confidence Interval) |
| --- | --- | --- | --- |
| Primary Composite Outcome | 332 (25.5%) | 859 (31.0%) | 0.82 (0.69-0.97) |
| Components of the Composite Outcome |  |  |  |
| All-cause Death | 203 (15.6%) | 608 (22.0%) | 0.82 (0.66-1.01) |
| Myocardial Infarction | 33 (2.5%) | 45 (1.6%) | 1.61 (0.88-2.93) |
| Ischemic Stroke | 12 (0.9%) | 50 (1.8%) | 0.68 (0.32-1.45) |
| Heart Failure | 115 (8.8%) | 244 (8.8%) | 0.78 (0.57-1.06) |
| Renal Failure | 26 (2.0%) | 77 (2.8%) | 0.57 (0.30-1.09) |
| Secondary Outcomes |  |  |  |
| Diabetic Ketoacidosis | 16 (1.2%) | 27 (0.9%) | 1.01 (0.42-2.44) |
| Severe Hypoglycemia | 10 (0.7%) | 35 (1.2%) | 1.08 (0.47-2.49) |
| 30-day Readmission | 174 (12.9%) | 488 (16.5%) | 0.73 (0.58-0.92) |

**Table 3.** Primary and secondary outcomes for SGLT2i compared to sulfonylurea.

| Outcome | SGLT2i | Sulfonylurea | Adjusted Hazard Ratio (95% Confidence Interval) |
| --- | --- | --- | --- |
| Primary Composite Outcome | 332 (25.5%) | 274 (26.0%) | 0.97 (0.80-1.18) |
| Components of the Composite |  |  |  |
| Death | 203 (15.6%) | 179 (17.0%) | 0.82 (0.66-1.01) |
| Myocardial Infarction | 33 (2.5%) | 15 (14%) | 1.41 (0.67-2.97) |
| Ischemic Stroke | 12 (0.9%) | 28 (2.7%) | 0.31 (0.14-1.72) |
| Heart Failure | 115 (8.8%) | 81 (7.7%) | 1.03 (0.72-1.46) |
| Renal Failure | 26 (2.0%) | 24 (2.3%) | 0.76 (0.38-1.50) |
| Secondary Outcomes |  |  |  |
| Diabetic Ketoacidosis | 16 (1.2%) | <6 (<1%) | 2.42 (0.66-8.85) |
| Severe Hypoglycemia | 10 (0.7%) | 9 (0.8%) | 0.75 (0.24-2.40) |
| 30-day Readmission | 174 (12.9%) | 180 (16.5%) | 0.66 (0.51-0.85) |

### Secondary outcomes

Prior to adjustment, the total number of patients with DKA was 16 (1.2%) in the SGLT2i group and 27 (0.9%) in the DPP4 group. After adjustment, this corresponded to a HR of 1.01 (95%CI 0.42-2.44). There was no significant difference in the rate of severe hypoglycemia between the SGLT2i and DPP4 groups (HR 1.08, 95% CI 0.47-2.49). 30-day readmission rates were lower in the SGLT2i compared to the DPP4 group (HR 0.73, 95%CI 0.58-0.92).

## Discussion

In this multicentre study of hospitalized older adults with type 2 diabetes mellitus, it was uncommon for patients to be newly started on an SGLT2i or GLP1a. New use of an SGLT2i, compared to a DPP4i, was associated with improved cardiovascular and renal outcomes, however the result was not robust in a sensitivity analysis where new use of a sulfonylurea was the comparator.

Multiple double-blind placebo-controlled randomized controlled studies have demonstrated that SGLT2i or GLP1a improve cardiovascular, heart failure, and renal outcomes.^31–33^ These results have also been replicated in observational studies of routine care. However, both the observational studies and randomized trials have predominantly included outpatients, rather than inpatients. This observation, coupled with editorials^34^, may explain why new use of SGLT2i and GLP1a remains low for hospitalized patients. This prevailing wisdom assumes that chronic conditions should not be managed in the inpatient setting and that the risks of starting either class of medications in the inpatient setting outweighs the potential benefits^35^. Results from our study suggest, relative to DPP4i, that SGLT2i are associated with improved cardiovascular and renal outcomes.

One of the few randomized trials of newly starting an SGLT2i in hospital was the EMPULSE trial^36^. It randomized adults with heart failure with or without diabetes to empagliflozin versus placebo and identified an improvement in the composite outcome of all-cause mortality, number and time to first HFEs, and change in Kansas City Cardiomyopathy Questionnaire Total Symptom Score (KCCQ-TSS) from baseline to 90 days (win ratio of 1.36 in favor of empagliflozin, 95% CI: 1.09–1.68). EMPULSE provides the highest quality data that the assumption SGLT2i should not be started in hospital is false. Other studies have shown that starting new medications for chronic diseases in-hospital, including at the time of discharge, leads to improved medication adherence compared to deferring initiation to the outpatient setting ^37–41^. For example, multiple qualitative studies of outpatient internists or family physicians have shown that they are more likely to continue medications started from a recent hospitalization, but are less likely to initiate a new prescription following a recent discharge from hospital ^42,43^.

There are also studies showing that newly starting medications for diabetes in the inpatient setting may not improve clinical outcomes. Specifically, Anderson et al. have shown that when patients were started on new or higher doses of diabetes medications, they had an increased risk of hypoglycemia with no improvement in hemoglobin A1c^44^. However, in their study, the vast majority of medication increases were for insulin or sulfonylureas. Both classes cause weight gain and hypoglycemia and do not improve cardiovascular outcomes.

Our study has several strengths. First, our cohort included linked inpatient and outpatient records. This provided a comprehensive dataset that included inpatient and outpatient medications, diagnoses, and investigations, and it allowed us to identify whether a medication had been newly initiated in the inpatient setting. One additional benefit of this linked dataset was the availability of hemoglobin A1c values. A recent inpatient cohort study from the GEMINI hospitals found that hemoglobin A1c was only available for roughly 20% of patients with diabetes^45^. In contrast, through linkage to outpatient laboratory tests, hemoglobin A1c was available for over 85% of the patients in our cohort. Second, our study predominantly focused on older adults, who are typically under-represented in randomized controlled trials^46,47^. Finally, we were able to assess for both effectiveness and safety outcomes, including diabetic ketoacidosis, which is a particular concern regarding the use of SGLT2i.

There are multiple limitations to our study. First, it is unclear why our results were not robust when sulfonylurea was the comparator group because sulfonylureas do not improve cardiovascular or renal outcomes. The null finding might reflect an indication bias. This is suggested by the fact that patients who received an SGLT2i, compared to sulfonylurea, had a higher prevalence of heart failure (37% vs 19%), coronary artery disease (19% vs 11%). Second, in Ontario, complete outpatient pharmacy data is only available within ICES for adults over the age of 65. Therefore, we restricted our study population to patients over 65 years of age, and how these results apply to younger adults is not known. Third, the outcomes of myocardial infarction, stroke, hospitalization for heart failure and renal failure were defined using ICD-10 codes. However, these codes have previously been shown to have high sensitivity and specificity.^48–51^ Additionally, use of ICD-10 codes should not have led to differential identification of outcomes between groups and therefore is unlikely to have resulted in measurement bias.

Adults with type 2 diabetes newly initiated on an SGLT2i during a hospitalization had a significantly reduced risk of major cardiovascular or renal events in the subsequent year compared to those initiated on a DPP4 inhibitor, but not compared to those initiated on a sulfonylurea. The prevalence of SGLT2i and GLP1a use among patients with an indication for these medications remains low. Future studies with a larger sample size are needed to better evaluate and characterize the potential risks and benefits of newly starting an SGLT2i or GLP1ra in hospital.

## Data Availability

All data produced in the present study are available upon submission of a project proposal to ICES in Ontario, Canada.

Study concept and design: All authors

Acquisition of data: All authors

Analysis/interpretation of data: All authors

Drafting of the manuscript: MC, MF

Critical revision of the manuscript: All authors

Statistical analysis: MH, MC, MF

## Acknowledgements

We thank IQVIA Solutions Canada Inc. for use of their Drug Information File. Parts of this material are based on data and/or information compiled and provided by CIHI and the Ontario Ministry of Health. The analyses, conclusions, opinions and statements expressed herein are solely those of the authors and do not reflect those of the funding or data sources; no endorsement is intended or should be inferred.

